# Abnormal Blood Pressure Dipping Pattern in Smokers

**DOI:** 10.1101/2025.02.03.25321626

**Authors:** Muhammed Ömer Arslan, Tijen Acar, Yunus Doğan, Şahbender Koç, İbrahim Sefa Güneş

## Abstract

**Aim:** Hypertension with non-dipper or reverse dipper patterns has worse outcomes. Smokers have some risk factors for abnormal dipping. There are conflicting results about the association between smoking and blood pressure (BP) dipping patterns, thus we aimed to examine it in essential hypertension.

**Methods:** 178 essential hypertension patients (ages 30-83) completed Fagerström Nicotine Dependence Test (FNDT) and our custom Patient Data Collection Test (PDCT). We analyzed these data with the ambulatory BP measurements (ABPM) and performed logistic regression analysis.

**Results:** Abnormal dipping patterns were significantly more frequent among patients with a history of smoking (75,0%) than those without history of smoking (47,4%), (p<0,001). Among smokers, abnormal dipping patients had significantly higher FNDT Scores (5 (0-10) versus 2 (0-9), p=0,046), longer smoking years (25 (6-50) versus 20 (5-50), p=0,017), and more smoking exposure in terms of pack.years (25 (3-135) versus 15 (1-75), p=0,023). Patients with history of smoking had significantly higher nocturnal systolic (128,10±15,54 versus 120,56±12,84 mmHg, p=0,001), nocturnal diastolic (80,82±12,60 versus 76,44±10,89 mmHg, p=0,016), and 24-hour mean systolic (133,15±13,52 versus 127,90±12,03 mmHg, p=0,008) BP values; and more blunted systolic and diastolic dipping ratios than non-smokers. Patients with a history of smoking were 3,484 (1,693-7,171 CI:0,95 p=0,001) times more likely to have abnormal dipping. Smoking was also associated with diabetes mellitus, dyslipidemia, and metabolic syndrome.

**Conclusion:** Smoking was associated with abnormal dipping patterns; higher nocturnal systolic, diastolic, and 24-hour systolic BP values; and lower systolic and diastolic dipping rates. FNDT score and smoking exposure parameters were significantly higher in abnormal dipping.

## Introduction

Hypertension is a major risk factor for cardiac disorders, stroke, renal diseases, dementia, and many other severe comorbidities.^(1-4)^ World Health Organization (WHO) estimates that globally 1,28 billion people 30 to 79 years of age have hypertension.^(5)^ It is actually the cause of death for 10,4 million people annually.^(6)^ Therapeutic guidelines of hypertension currently focus on average Blood Pressure (BP) levels.^(1-4)^ However, research points out the importance of particular sequential BP values and BP fluctuations beyond average BP values.^(7, 8)^

BP is not a static physical feature; instead, it is a highly dynamic biological aspect of life exhibiting regional and temporal variability in the body. As the body has two diurnal phases, one awake and the other one asleep, it tries to maintain two different levels of dynamic stability, with these two periods having their own normals.^(2-4)^ Thus, BP exhibits a diurnal variation, and normally declines during sleeping by 10% to 20% compared to the awake period.^(9, 10)^

Normal BP dipping phenomenon reflects an end result of the normal functioning of circadian rhythm, water and sodium regulation, and multiple interplaying neuroendocrine mechanisms.^(7, 11)^ On the other hand, genetic and environmental factors may influence the BP dipping pattern.^(11)^ In fact, deviant diurnal variations are shown to be associated with an increased risk of diabetes mellitus, cardiovascular autonomic neuropathy, and overall cardiovascular morbidity and mortality.^(12)^

Abnormal dipping patterns are extreme dipper, non-dipper, and reverse dipper patterns. If the BP decline is above 20%, it is called as extreme dipper pattern. In the case of a non-dipper pattern, BP declines between 0-10% compared to the awake period. The reverse dipper pattern exhibits a rise in BP during sleeping compared to the awake period.^(11, 12)^ These phenomena occur both in normotensive and hypertensive individuals.^(13)^ Hypertension types with non-dipper or reverse dipper patterns of BP are associated with increased cardiovascular and overall morbidity and mortality^(10, 12)^, increased risk of stroke^(10)^, heart failure^(14)^, cognitive impairment, and dementia.^(15)^ Furthermore, DNA damage markers that are shown in hypertensive patients are higher in non-dipper hypertension.^(16, 17)^

On the other hand, smoking is a major causative risk factor for a variety of comorbidities like many types of cancers^(18, 19)^, chronic obstructive pulmonary disease^(20)^, coronary heart disease, atherosclerosis, cerebrovascular disease, and hypertension^(4, 21)^. Nevertheless, due to nicotine, tobacco smoking is highly addictive and, thus, comprises another serious and widespread concern of public health.^(22, 23)^ According to WHO data, 22,3% of the total world population, 36,7% of men and 7,8% of women smoke.^(24)^ Currently, over 8 million deaths annually are linked to smoking, of which 1.3 million are attributed to secondhand exposure.^(24)^

As abnormal dipping patterns are associated with increased morbidity and mortality^(9-14)^, it is critical to know if there are further associations between smoking and abnormal dipping patterns, which may shed light on the effects of smoking and how to tackle them. Some studies identify a correlation between smoking and hypertension with non-dipper and reverse dipper patterns.^(25, 26)^ Others relate no such correlation^(27-32)^; some even indicate a negative correlation.^(13, 33-36)^ There are some discrepancies regarding the definitions and classifications of BP dipping patterns, the inclusion criteria of the studies, and the grouping of the patients among these studies, which may account for the conflicting results.^(11)^ Therefore, we have primarily aimed to achieve a consistent and clear view regarding the particular relationship between tobacco smoking and abnormal BP dipping in essential hypertension.

## Methods

### Study Design and Population

Our study was a multidisciplinary prospective case-control study in Ankara Sanatoryum Research and Education Hospital (ASREH), conducted with 178 essential hypertension patients who were referred to 24-hour Ambulatory Blood Pressure Measurement (ABPM) by their primary physicians. Hypertension was defined according to the 2017 AHA/AAC^(3)^, 2018 ESC^(4)^, and 2020 ISH^(2)^ Guidelines. This study was conducted with those eligible patients (n=178) applying consecutively to the ABPM Test Unit of the hospital between the dates of 15.06.2023 and 15.08.2023 and who were willing to participate in our study with their informed consent.

Exclusion criteria were having a secondary hypertension diagnosis, or a diagnosis of a sleep disorder, describing an inability to sleep well during the test, and being under the age of 30.^(11, 37, 38)^ As secondary hypertension causes abnormal dipping patterns and may interfere with the results, it is excluded in order to demonstrate exactly the sole effect of smoking on the BP dipping pattern.^(11)^ Hypertension under the age of 30 grounds a suspicion of secondary hypertension, and some authors recommend a thorough search for such etiologies.^(37, 39)^ For that reason, we excluded such patients too.

Patients who had more than 20% unsuccessful measurements during the awake-time or had less than 4 successful measurements during sleeping were also excluded from the study.^(40)^ We did not accept a standardized day and night time separation; instead, we questioned each participant’s awake and sleeping times and calculated their awake and sleeping average BP values accordingly. In concordance with most previous studies, systolic BP measurements were utilized to categorize the type of BP dipping pattern of the patients.^(10)^ Each consecutive patient was thoroughly evaluated when they completed the ABPM and came to return the measurement device, and if eligible, the patient was included in our study. We have accepted the dipper group as our control group. Extreme dipper, non-dipper, and reverse dipper patterns were categorized together as abnormal dipping patterns.

This study complies with the Declaration of Helsinki on the ethical principles for medical research. The scientific monitoring board of ASREH approved the study design.

### Data Sources

Patients were questioned using our custom Patient Data Collection Test (PDCT) and Fagerström Nicotine Dependency Test (FNDT).^(41-43)^ PDCT documented the variables of age, sex, height, weight, body mass index (BMI), waist and neck circumference, comorbidities, antihypertensive drug use, if using any antihypertensives then the timing of the drugs, and smoking status (never smoked, quit, smoking actively). Anamnesis, past medical records, and test results of each patient were evaluated carefully for comorbidities. If the patient had a history of smoking (either actively or formerly), then it was further detailed concerning the years of smoking and amounts of daily smoking. The minimal abstinence cutoff was accepted as six months in order to classify a patient as quit.^(44)^

Our patients defined their tobacco use exclusively as cigarette smoking. We quantified the amount of smoking as pack.years, which is the multiplication of years of smoking with the average amount of packs consumed per day. Each pack contains 20 cigarettes in Türkiye.

As for the antihypertensive use, we counted each antihypertensive substance, not the pills used. Antihypertensive timing is categorized as morning, noon, night, and other possible combinations.

FNDT is a standardized 6-question test for the objective and quantitative assessment of nicotine dependence.^(41, 42)^ Validated Turkish version^(43)^ of the FNDT was used for the questionnaire of active smokers.

ABPMs were taken with SunTech Bravo 24H ABP device. Awake-time measurements were taken every half an hour, and sleep-time measurements were taken hourly. ABPM results were evaluated together with the data acquired from questionnaires. ABPM records documented average awake-time BP values, sleep-time BP values in mmHg, and BP dipping values in percentage (%).

### Statistical Methods

The conformity of the variables to normal distribution was determined by Shapiro-Wilk and Kolmogorov-Smirnov tests, skewness and kurtosis values, histogram graphs, mean ± standard deviation, and median values. Numerical variables that fit the normal distribution are expressed as mean ± standard deviation, and numerical variables that do not fit the normal distribution are expressed as median (min-max). Categorical variables were expressed as numbers (percentage distributions). Student T-test was used for comparisons between independent groups for continuous numerical variables that fit the normal distribution. Mann-Whitney U test was used for comparisons between independent groups for continuous numerical variables that do not fit the normal distribution. Pearson Chi-square test, Fisher’s Exact Test, and Yates corrected chi-square test were used for intergroup comparisons of categorical variables.

Multivariate logistic regression analysis was performed by constituting a model with variables that were considered clinically important for dipping patterns, and odds ratio values were calculated.

A value of p<0.05 was considered statistically significant in the analysis of all tests, and these analyses were performed using the SPSS 25.0 program.

## Results

The patients (n=178) were aged between 30-83 years old and consisted of 84 (47,2%) females and 94 (52,8%) males (**Table 1**). The average age of male and female participants was similar: 54,33±10,61 years for females, and 54,69±12,20 years for males (p=0,711). Anthropometric measurements of patients were distributed normally. The average values were 165,30±9,65 cm for height; 80,41±15,18 kg for weight; 29,35±4,81 kg/m2 for BMI; 99,63±12,68 cm for waist circumference; and 38,69±3,72 cm for neck circumference. BMI values were similar between male and female groups. 78 (43,8%) patients have never smoked, 37 (20,8%) patients had smoked but quit, and 63 (35,4%) patients have smoked actively. Median smoking years among patients with a history of smoking (active or quit; n=100) were 25 (6-50) years. The median smoking amount in terms of pack.years was 23 (1-135). The median time of abstinence among those defined as quit was 15 (1-43) years.

**Table 1.** Main Characteristics of the Study Population (n=178)

| Characteristics | Study Population (n=178) |
| --- | --- |
| Age (years) | 54,63±11,36 |
| Sex (female) | 84 (47,2) |
| Height (cm) | 165,30±9,65 |
| Weight (kg) | 80,41±15,18 |
| BMI (kg/m<sup>2</sup>) | 29,35±4,81 |
| Waist Circumference (cm) | 99,63±12,68 |
| Neck Circumference (cm) | 38,69±3,72 |
| Comorbidities: |  |
| DM | 50 (28,1) |
| Prediabetes | 15 (8,4) |
| DL | 101 (56,7) |
| MetS | 102 (57,3) |
| CAD and/or HF | 18 (10,1) |
| Asthma and/or COPD | 10 (5,6) |
| CKD | 6 (3,4) |
| Tobacco Smoking: |  |
| Never Smoked | 78 (43,8) |
| Quit | 37 (20,8) |
| Actively Smoking | 63 (35,4) |
| Smoking Years | 25 (6-50) |
| Smoking pack.years | 23 (1-135) |
| Abstinence Years (n=37) | 15 (1-43) |
| Total FNDT Score (n=63) | 5 (0-10) |
| BP Dipping Pattern |  |
| Dipper | 66 (37,1) |
| Extreme Dipper | 3 (1,7) |
| Non-dipper | 76 (42,7) |
| Reverse Dipper | 33 (18,5) |
| Isolated Nocturnal HT | 29 (16,3) |
| Anti-HT Use | 102 (57,3) |
| Anti-HT Count | 2 (1-5) |
| 1 | 32 (31,4) |
| 2 | 44 (43,1) |
| 3 | 21 (20,6) |
| 4 | 4 (3,9) |
| 5 | 1 (1,0) |
| Anti-HT Timing |  |
| Morning | 75 (73,5) |
| Noon | 1 (1,0) |
| Night | 5 (4,9) |
| Morning and Noon | 2 (2,0) |
| Morning and Night | 13 (12,7) |
| Morning, Noon and Night | 6 (5,9) |
| HT Diagnosis |  |
| New Diagnosed | 77 (43,3) |
| Diagnosed Before | 101 (56,7) |
| BP Values (mmHg) |  |
| 24-h Mean SBP | 130,85±13,12 |
| 24-h Mean DBP | 83,62±10,40 |
| Awake Mean SBP | 133,34±13,33 |
| Awake Mean DBP | 85,51±10,54 |
| Sleeping Mean SBP | 124,80±14,86 |
| Sleeping Mean DBP | 78,90±12,05 |
| Systolic Dipping (%) | 6 ((-11)- 22) |
| Diastolic Dipping (%) | 7 ((-11)- 36) |

Data are expressed as mean±ss, median (min-max), and n (%). anti-HT: antihypertensive, BP: Blood Pressure, BMI: body mass index, CAD: Coronary Artery Disease, CKD: Chronic Kidney Disease, COPD: Chronic Obstructive Pulmonary Disease, DBP: Diastolic Blood Pressure, DM: Diabetes Mellitus, DL: Dyslipidemia, FNDT: Fagerström Nicotine Dependence Test, HF: Heart Failure, HT: hypertension, MetS: Metabolic Syndrome, SBP: Systolic Blood Pressure

There were 66 (37,1%) dipper, 3 (1,7%) extreme dipper, 76 (42,7%) non-dipper, and 33 (18,5%) reverse dipper patients. 77 (43,3%) of the study population were newly diagnosed with the current ABPM, and 101 (56,7%) of them were already diagnosed with essential hypertension. 102 (57,3%) patients were under medical antihypertensive treatment. Median number of antihypertensives used was 2 (1-5). Newly diagnosed patients’ distribution, antihypertensive use status, and antihypertensive substance counts were similar between the female and male groups. The median score of FNDT was 5 (0-10). There were 30,2% very low; 19,1% low; 7,9% medium; 22,2% high; 20,6% very high degrees of nicotine dependence among smokers. Patient population characteristics are further detailed in **Table 1**.

Patient characteristics according to BP dipping patterns are detailed in **Table 2**. There were no significant differences between the normal dipper group and abnormal dipping group with regard to their anthropometric specifications, comorbidities, time of hypertension diagnosis (new versus established earlier), antihypertensive use (present versus absent), antihypertensive count and antihypertensive timing (night versus others). Remarkably, we also checked for an association between abnormal dipping and waist or neck circumference threshold values; however, we did not find significant differences. Dipper BP rates (52,6%) in patients without any history of smoking (n:78) were significantly higher than the rates of dipper BP (25,0%) in the patients with a history of smoking (active or past) (n:100), (p<0,001). Overall, abnormal BP dipping patterns were significantly more frequent among patients with a history of smoking (75,0%) than among those without any history of smoking (47,4%), (p<0,001). Logistic regression analysis showed that patients with a history of smoking were 3,484 (1,693-7,171 CI:0,95 p=0,001) times more likely to have an abnormal dipping pattern (**Table 4**).

**Table 2.** Distribution of Main Characteristics According to BP Dipping Pattern.

| Variable | Normal | Abnormal dipping patterns |  |  |  | p-values<br>dipper vs<br>abnormal |
| --- | --- | --- | --- | --- | --- | --- |
|  | Dipper<br>66 (37,1) | Extreme dipper<br>3 (1,7) | Non-dipper<br>76 (42,7) | Reverse dipper<br>33 (18,5) | All<br>Abnormal<br>Dippers<br>112 (62,9) |  |
| Sex, n (%) female | 35 (37,2) | 2 (2,1) | 40 (42,6) | 17 (18,1) | 59 (62,8) | 0,964<sup>1</sup> |
| Age (years) | 53,50±12,37 | 49,33±1,15 | 53,78±10,71 | 59,33±10,23 | 55,29±10,73 | 0,310<sup>2</sup> |
| Height (cm) | 164,91±9,75 | 169,33±15,04 | 165,38±9,01 | 165,52±10,75 | 165,53±9,63 | 0,681<sup>2</sup> |
| Weight (kg) | 81,15±14,84 | 83,00±20,52 | 79,75±15,59 | 80,21±15,11 | 79,97±15,43 | 0,618<sup>2</sup> |
| BMI (kg/m<sup>2</sup>) | 29,84±4,67 | 28,57±2,15 | 29,03±5,04 | 29,21±4,78 | 29,07±4,88 | 0,304<sup>2</sup> |
| Waist circumference<br>(cm) | 99,76±11,74 | 90,33±9,50 | 99,68±13,64 | 100,09±12,66 | 99,55±13,26 | 0,918<sup>2</sup> |
| Neck circumference<br>(cm) | 38,83±3,67 | 39,33±4,93 | 38,85±3,64 | 37,97±3,98 | 38,60±3,76 | 0,690<sup>2</sup> |
| BMI >30 kg/m<sup>2</sup> | 30 (43,5) | 1 (1,4) | 28 (40,6) | 10 (14,5) | 39 (56,5) | 0,160<sup>1</sup> |
| Waist c. >100 cm in m.,<br>>90 cm in f. | 43 (38,1) | 0 (0,0) | 50 (44,2) | 33 (17,7) | 83 (61,9) | 0,846<sup>3</sup> |
| Neck c. >43 cm in m.,<br>41 cm. in f. | 10 (45,5) | 1 (4,5) | 8 (36,4) | 3 (13,6) | 12 (54,5) | 0,527<sup>3</sup> |
| Comorbidities, n (%) |  |  |  |  |  |  |
| DM | 14 (28,0) | 1 (2,0) | 22 (44,0) | 13 (26,0) | 36 (72,0) | 0,163<sup>3</sup> |
| Prediabetes | 7 (46,7) | 1 (6,6) | 3 (20,0) | 4 (26,7) | 8 (53,3) | 0,600<sup>3</sup> |
| MetS | 34 (33,3) | 2 (2,0) | 47 (46,1) | 19 (18,6) | 68 (66,7) | 0,231<sup>1</sup> |
| DL | 34 (33,7) | 2 (2,0) | 48 (47,5) | 17 (16,8) | 67 (66,3) | 0,280<sup>1</sup> |
| CAD/HF | 7 (38,9) | 0 (0,0) | 9 (50,0) | 2 (11,1) | 11 (61,1) | 1,000<sup>3</sup> |
| Asthma/COPD | 4 (40,0) | 0 (0,0) | 3 (30,0) | 3 (30,0) | 6 (60,0) | 1,000<sup>4</sup> |
| CKD | 2 (33,3) | 0 (0,0) | 2 (33,3) | 2 (33,3) | 4 (66,7) | 1,000<sup>4</sup> |
| Smoking Status, n (%) |  |  |  |  |  |  |
| Never Smoked | 41 (52,6) | 1 (1,3) | 27 (34,6) | 9 (11,5) | 37 (47,4) | **0,001<sup>1</sup>** |
| Quit | 9 (24,3) | 1 (2,7) | 19 (51,4) | 8 (21,6) | 28 (75,7) |  |
| Actively Smoking | 16 (25,4) | 1 (1,6) | 30 (47,6) | 16 (25,4) | 47 (74,6) |  |
| Smoking History, n<br>(%) |  |  |  |  |  |  |
| Never Smoked | 41 (52,6) | 1 (1,3) | 27 (34,6) | 9 (11,5) | 37 (47,4) | **<0,001<sup>1</sup>** |
| Smoked (active or<br>quit) | 25 (25,0) | 2 (2,0) | 49 (49,0) | 24 (24,0) | 75 (75,0) |  |
| Smoking Years (n=100) | 20 (7-50) | 10 (10-10) | 25 (6-50) | 27 (13-50) | 25 (6-50) | **0,017<sup>5</sup>** |
| Pack.years (n=100) | 15 (1-75) | 4 (3-5) | 25 (4-135) | 25 (4-135) | 25 (3-135) | **0,023<sup>5</sup>** |
| FNDT Total Score | 2 (0-9) | 4 (4-4) | 6 (0-10) | 4 (0-10) | 5 (0-10) | **0,046<sup>5</sup>** |
| (n=78) |  |  |  |  |  |  |
| 1. Question <2 points | 5 (13,5) | 1 (2,7) | 21 (56,8) | 10 (27,0) | 32 (86,5) | 0,022<sup>3</sup> |
| 1. Question ≥2 points | 11 (42,3) | 0 (0,0) | 9 (34,6) | 6 (23,1) | 15 (57,7) |  |
| 2. Question 0 points | 13 (35,1) | 1 (2,7) | 14 (37,8) | 9 (24,3) | 24 (64,9) | 0,068<sup>3</sup> |
| 2. Question 1 points | 3 (11,5) | 0 (0,0) | 16 (61,5) | 7 (26,9) | 23 (88,5) |  |
| Anti-HT Use, active | 35 (34,3) | 0 (0,0) | 46 (45,1) | 21 (20,6) | 63 (65,7) | 0,376<sup>1</sup> |
| Anti-HT Count (n=102) | 2 (1-5) | - | 2 (1-4) | 1 (0-4) | 2 (1-4) | 0,161<sup>5</sup> |
| Isolated Nocturnal Hypertension, n (%) | 4 (13,8) | 0 (0,0) | 14 (48,3) | 11 (37,9) | 25 (86,2) | 0,009<sup>3</sup> |
| Anti-HT Timing (n=102) |  |  |  |  |  |  |
| Night | 8 (33,3) | 0 (0,0) | 11 (45,8) | 5 (20,8) | 16 (66,7) | 1,000<sup>3</sup> |
| Other than Night | 27 (34,6) | 0 (0,0) | 35 (44,9) | 16 (20,5) | 51 (65,4) |  |
| HT Diagnosis |  |  |  |  |  |  |
| New Diagnosed | 33 (42,8) | 3 (3,9) | 29 (37,7) | 12 (15,6) | 44 (57,1) | 0,163<sup>1</sup> |
| Diagnosed Before | 33 (32,7) | 0 (0,0) | 47 (46,5) | 21 (20,8) | 68 (67,3) |  |
| BP Values (mmHg) |  |  |  |  |  |  |
| 24-h Mean SBP | 129,41±11,54 | 134,33±3,06 | 129,79±13,36 | 135,85±15,12 | 131,70±13,95 | 0,262<sup>2</sup> |
| 24-h Mean DBP | 82,41±9,53 | 79,67±10,02 | 83,25±10,14 | 87,24±12,18 | 84,33±10,86 | 0,235<sup>2</sup> |
| Awake Mean SBP | 134,50±12,03 | 142,67±3,51 | 131,49±13,68 | 134,45±15,13 | 132,66±14,05 | 0,375<sup>2</sup> |
| Awake Mean DBP | 86,14±10,02 | 84,33±13,01 | 84,59±10,40 | 86,45±11,91 | 85,13±10,85 | 0,541<sup>2</sup> |
| Sleeping Mean SBP | 117,27±10,30 | 111,67±4,04 | 125,64±13,34 | 139,09±15,57 | 129,23±15,38 | <0,001<sup>2</sup> |
| Sleeping Mean DBP | 73,48±9,51 | 67,33±6,03 | 79,84±10,79 | 88,61±13,10 | 82,09±12,28 | <0,001<sup>2</sup> |

<sup>1</sup>Pearson Chi-Square test, <sup>2</sup>Student t test, <sup>3</sup>Yates corrected chi-square test, <sup>4</sup>Fisher's Exact test, <sup>5</sup>Mann-Whitney U test; Data are expressed as mean±ss, median (min-max) and n (%). Row percentages are given. anti-HT: antihypertensive, BP: Blood Pressure, BMI: body mass index, CAD: Coronary Artery Disease, CKD: Chronic Kidney Disease, COPD: Chronic Obstructive Pulmonary Disease, DBP: Diastolic Blood Pressure, DM: Diabetes Mellitus, DL: Dyslipidemia, FNDT: Fagerström Nicotine Dependence Test, HF: Heart Failure, HT: hypertension, MetS: Metabolic Syndrome, SBP: Systolic Blood Pressure

**Table 3.**
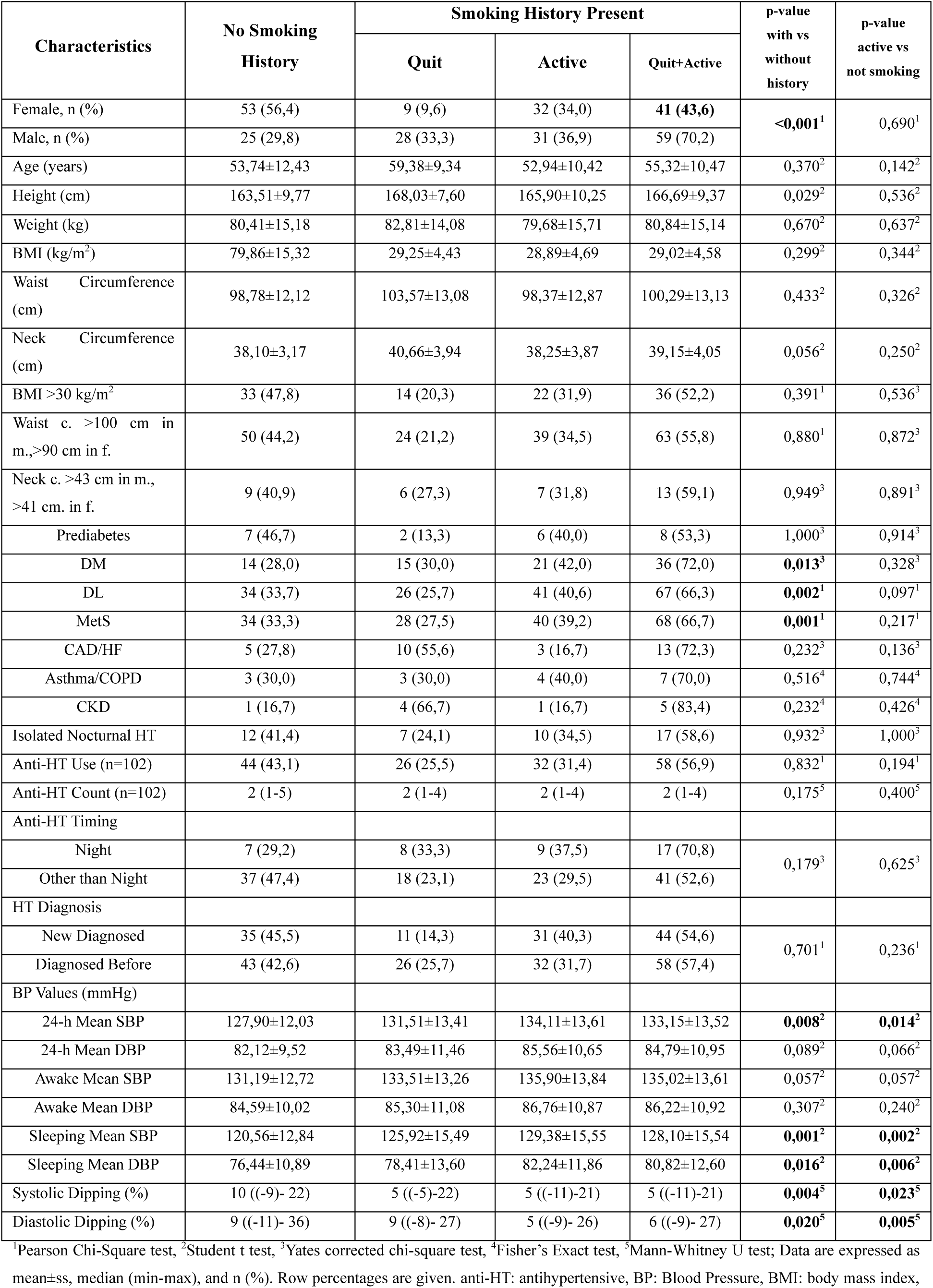
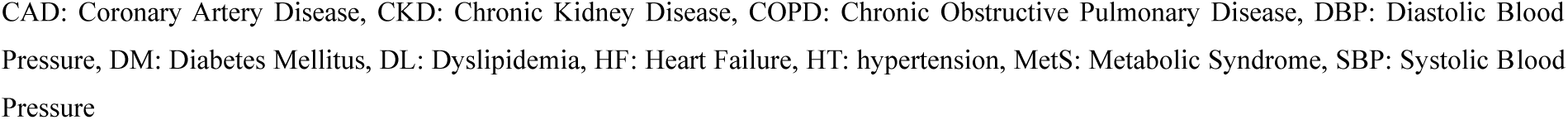
Main Characteristics of the Study Population According to Tobacco Smoking Status(n=178)

**Table 4.** Results of multivariate logistic regression analysis for abnormal dipping *.

|  | **OR (%95 CI)** | **p-value** |
| --- | --- | --- |
| Age | 0,999 (0,968-1,032) | 0,963 |
| Male (ref: Female) | 0,879 (0,336-2,300) | 0,793 |
| BMI | 0,968 (0,891-1,053) | 0,452 |
| Neck Circumference | 0,974 (0,847-1,119) | 0,710 |
| DM | 1,554 (0,642-3,762) | 0,328 |
| DL | 1,048 (0,447-2,454) | 0,914 |
| MetS | 0,887 (0,339-2,320) | 0,806 |
| Isolated Nocturnal HT | **4,905** (1,502-16,019) | **0,008** |
| Diagnosed Before HT (ref: newly diagnosed HT) | 1,571 (0,492-5,023) | 0,446 |
| Anti-HT Medication Number | 0,967 (0,604-1,550) | 0,890 |
| Smoking Tobacco | **3,514** (1,712-7,211) | **0,001** |

\*Hosmer and Lemeshow test=0,977, Nagelkerke R<sup>2</sup>=0,196
CI: confidence interval, OR: odds ratio, anti-HT: antihypertensive, BMI: body mass index, DM: Diabetes Mellitus, DL: Dyslipidemia, HT: hypertension, MetS: Metabolic Syndrome

Abnormal dipping patients had significantly higher FNDT Scores (5 (0-10) versus 2 (0-9), p=0,046), longer smoking years (25 (6-50) versus 20 (5-50), p=0,017), and more tobacco smoking exposure in terms of pack.years (25 (3-135) versus 15 (1-75), p=0,023). More specifically, actively smoking and abnormal dipping BP patients were significantly more likely to score equal to or higher than 2 points in the first question (which questions the timing of first cigarettes upon awakening) of FNDT compared to dipper patients, which in turn indicates some further association between more specific traits of nicotine dependence and abnormal dipping BP pattern (**Table 2)**.

Abnormal dipping patients had also higher rates of isolated nocturnal HT (86,2% versus 13,8%, p=0,009) and higher mean nocturnal systolic (129,23±15,38 mmHg versus 117,27±10,30 mmHg, p<0,001) and diastolic (82,09±12,28 mmHg versus 73,48±9,51 mmHg, p<0,001) BP values. 24-hour and awake-time averages of both systolic and diastolic BP levels did not differ significantly (**Table 2**). Isolated nocturnal hypertension patients (patients with hypertension during sleeping but normal BP levels during awake period) were significantly at greater risk for abnormal dipping BP patterns in the multivariate logistic regression analysis; OR= 4,905( CI: 1,502-16,019) (**Table 4**). Isolated nocturnal hypertension is defined as hypertension with normal daytime and office BP.^(9)^ Nocturnal hypertension is not synonymous with non-dipper or reverse dipper hypertension.^(9)^ Patients with normal dipper patterns may have nocturnal hypertension.^(45)^

We have found that patients with a history of smoking had significantly higher nocturnal systolic (128,10±15,54 versus 120,56±12,84 mmHg, p=0,001), nocturnal diastolic (80,82±12,60 versus 76,44±10,89 mmHg, p=0,016), and 24-hour mean systolic (133,15±13,52 versus 127,90±12,03 mmHg, p=0,008) BP values; and more blunted systolic (5 ((-11)-21) % versus 10 ((-9)-22) %, p=0,004) and diastolic (6 ((-9)-27) % versus 9 ((-11)-36) %, p=0,020) dipping ratios than those patients without any history of smoking (**Table 3**).

History of smoking was associated significantly with being male, having diabetes mellitus, dyslipidemia, and metabolic syndrome (**Table 3**). Remarkably, this association was detected between history of smoking and non-smokers; however, it was not detected between active smoking and not smoking actively (ie quit and never-smokers together).

## Discussion

In this study, patients with a history of smoking showed significantly more blunted dipping patterns both for systolic BP [5 ((-11)-22)% for smokers versus 10 ((-9)-22) % for non-smokers, p=0,004] and diastolic BP [6 ((-9)-27) % for smokers versus 9 ((-11)-36) % for non-smokers, p=0,020]. Patients with a history of smoking were 3,484 (1,693-7,171 CI:0,95 p=0,001) times more likely to have an abnormal dipping pattern (**Table 4**). Age, sex, neck circumference, and BMI showed no significant differences in regression analysis. These parameters were similar between our groups. Other studies reported significantly higher age^(33, 35)^ and BMI^(33)^ values for non-dipping groups. However, we did not detect such differences possibly because of homogeneity in our study population. This homogeneity in terms of anthropometric values may help us observe the effect of other variables (eg. smoking) upon BP dipping pattern.

Some studies reported contrasting results with our findings, e.g., Hermida et al^(13)^, de la Sierra et al^(33)^, Kaya MG et al^(34)^, Chotruangnapa et al^(35)^, Boggia et al.^(36)^ In fact, these studies reported an inverse relationship between smoking and non-dipping in their study groups. However, there were a number of important limitations in those studies. For example, although Hermida et al had accepted the diagnosis of secondary hypertension as an exclusion criterion, they did not exclude obstructive sleep apnea patients.^(13)^ Additionally, their non-dipper group had been significantly older, which could have affected the results because it was reported that the non-dipper pattern increased with aging.^(25)^ Furthermore, they included only patients under treatment with multiagent antihypertensives, which in turn could bias the results. Lastly, they had exercised their study with 48 hours of ABPM, and this relatively long testing time might have caused discomfort and stress in patients, which in turn might have influenced and masked the actual dipping pattern of the patients.^(38, 46)^

The study of de la Sierra et al was retrospective, and they did not report exclusion of secondary hypertension patients.^(33)^ In addition, the retrospective character of the study makes it very hard to evaluate the sleep quality of the patients. Also, they reported significant differences between their study groups concerning comorbidities and age, which may have affected the results.^(25)^ Again, Kaya MG et al^(34)^, Chotruangnapa et al^(35)^, and Boggia et al^(36)^ did not report any explicit effort to exclude secondary hypertension patients and sleep disturbances, making the results of their studies problematic from our perspective.^(11)^ In addition, the retrospective nature of the study of Chotruangnapa et al^(35)^ brings about uncertainty about the evaluation of sleep disturbances and the smoking status of patients. Clinical records about smoking may be incomplete because they are taken with different intentions, and sometimes, smoking may not be the primary concern at the clinical meeting. Hence, retrospective studies about smoking have practical limitations. Again, Boggia et al also reported significant differences in age and antihypertensive use between dipping groups.^(36)^

Borrelli et al^(27)^ reported no significant association between smoking status and dipping status of the patients. However, their study was performed in a CKD setting which has a very different nature in itself compared to essential hypertension, that is determining the patterns of patients’ BP. They reported significant differences in GFR levels of their patients. Besides, they did not exclude other types of secondary hypertension. These factors might have concealed the effect of smoking itself.^(11, 27)^

Gunebakmaz et al^(28)^, Ozcan et al^(30)^, Sunbul et al^(31)^, and Demir^(32)^ have also reported no significant difference. However, these studies were retrospective, and their hypertensive patient groups were heterogeneous, with no report of exclusion of secondary hypertension or sleep disturbances. The heterogeneity of their groups might have concealed the effect of smoking on dipping patterns. Palatini et al^(29)^ also reported an absence of association between smoking and non-dipping. However, their study did not report any exclusion of sleep disturbances, which might have affected their results.^(11)^

Brotman et al^(25)^ and Cai et al^(26)^ reported results similar to our study regarding the association between smoking and non-dipping BP patterns. However, their studies were retrospective, and secondary hypertension patients were not excluded, which caused uncertainty about their results.^(11)^

Our findings suggest a link between abnormal dipping status and nicotine dependency beyond smoking per se. According to our results, higher FNDT scores (5 (0-10) versus 2 (0-9), p=0,046), longer smoking years (25 (6-50) versus 20 (5-50), p=0,017), and more tobacco smoking exposure in terms of pack.years (25 (3-135) versus 15 (1-75), p=0,023) were associated with abnormal BP dipping patterns. To our knowledge, there have been no studies evaluating the relationship between nicotine dependency and BP dipping patterns. This result invites further research on this topic. The higher scores on the first question of the FNDT and the higher total FNDT scores or higher smoking exposure should prompt an ABPM of the patient to check for any nocturnal hypertension or abnormal dipping status.

Smokers have a relatively long nicotine-free period during sleeping. Patients reported even awakening during the night due to nicotine cravings in a study.^(47)^ Nicotine deprivation causes stress and increases the stress-related hormones and neurotransmitters. The noradrenergic system is activated and reinforces the craving symptoms.^(48)^ These neurohormonal changes due to nicotine deprivation may be a possible mechanism explaining the increase in nocturnal BP values and blunted BP dipping status in our study. One study found that alpha blockage restored the normal dipping pattern.^(49)^ Another study found that alpha blockage assisted patients in coping with nicotine deprivation symptoms.^(48)^ Our findings are in line with this evidence, suggesting a link between higher nicotine dependence, ie, higher FNDT scores and higher rates of nocturnal hypertension, non-dipper, and reverse dipper patterns. Other studies reported melatonin deficiency in smoking groups.^(50)^ Melatonin is an important factor in maintaining normal diurnal rhythm. Melatonin deficiency may be another mechanism behind the abnormal dipping.

We observed that there was a link between smoking and isolated nocturnal hypertension, higher 24-hour systolic, nocturnal systolic, and nocturnal diastolic BP levels in our study population. Isolated nocturnal hypertension and nocturnal hypertension patients were shown to be at increased risk for target organ damage due to hypertension, and their risks for cardiovascular events and mortality were higher.^(9, 45, 51, 52)^

There is evidence to recommend the treatment of isolated nocturnal hypertension; otherwise, these patients would be at higher risk of cardiovascular morbidity and mortality.^(53, 54)^ Thus, it is of note that clinicians should be aware of the probability that a smoker may present with nocturnal hypertension despite being normotensive in office BP measurements. Due to their increased risk of nocturnal hypertension, patients with a history of smoking may be at higher risk for cardiovascular events and mortalities, even if they show normotensive values in office or daytime home BP measurements. It is unpractical to perform ABPM for general population screening purposes; however, performing ABPM on selected at-risk patients might be of benefit with applicability also. History of smoking may be proposed as a criterion of predictive value for determining such at-risk patients.

We did not find any association between the dipping status of the patients and whether the patients took their antihypertensive medications at night or not. HYGIA and MAPEC trials, among many other studies, have found that chronotherapy or administering antihypertensive medications at night was effective for reestablishing the normal dipping status.^(55-57)^ These trials also reported significant cardiovascular benefits with night administration of antihypertensive medications.^(55-57)^ However, TIME study found that there was no significant change in cardiovascular adverse event rates with the ingestion of the antihypertensive medication at night.^(58, 59)^ Additionally HARMONY trial laid proof that there were no significant changes in dipping pattern or cardiovascular events upon nighttime administering of antihypertensive medications.^(60)^ Our findings are in line with the results of HARMONY trial. Antihypertensive drug amounts in the blood reach a plateau level after a period of regular use, which may explain the indifference in effects.^(61)^

Our findings agreed with the majority of the current scientific research in pointing out the association of a history of smoking with diabetes mellitus^(62, 63)^ and dyslipidemia^(63, 64)^. Visceral obesity, which is a strong indicator of metabolic syndrome, is increased in smoking patients.^(62)^ A meta-analysis found that MetS was increased with smoking.^(65)^ Our findings demonstrated an increased rate of MetS in the groups with a history of smoking; however, we did not find any significant difference neither in waist circumference nor in neck circumference as continuous variables and as variables with cutoff values. The main association was between history of smoking and non-smokers; however, it was not detected between active smoking and not smoking actively (ie quit and never-smokers together). This suggests a remaining link between smoking and comorbidities even after quit.

## Limitations

Despite the fact that many automatic devices that are used for ABPM can give false results if the patient has cardiac arrhythmias, we and many other researchers didn’t exclude those patients. The number of such patients was 6 in our study, so we have deemed this relatively small number as not changing the results. However, it should be reported as a limitation of our study. In addition, there was an insufficient number of extreme dipper patients (n=3), which prevented us from reaching any significant conclusions about this condition. Our classifications rested upon only one ABPM test, but research suggests some limitations regarding the reproducibility of BP dipping patterns on repeat testing at individual level.^(46)^ Thus, our one-time testing should be noted as another limitation of our study. More experimental and observatory studies with longitudinal follow-up and repeat testing plans are needed. Nevertheless, at the population level, ABPM has been shown to hold a valuable consistency with similar results on the overall ratios.^(46)^ Therefore, even one-time testing preserves an important significance for the population level. Additionally, there is a need to implement a broader multidisciplinary team approach to avoid positive outcome bias in further studies. Furthermore, ESC guidelines recommend evaluation for secondary hypertension under the age of 40.^(4)^ Our age cutoff of 30 might have brought about letting in some more secondary hypertension patients than a cutoff of 40 years of age. In addition, we did not perform polysomnography, renal ultrasonography, or endocrinological blood tests such as renin, aldosterone, or cortisol. We relied on the current results and diagnosis of the patients. Absolute exclusion of secondary hypertension patients might have not been achieved. Another limitation of our study was not describing antihypertensives in detail and taking each one as equals, however, evidence suggests there are differences in effect for dipping among anti-hypertensive medications.^(11)^

## Conclusion

Smoking was associated with more blunted diurnal systolic and diastolic BP dipping levels, in addition to its association with higher absolute values of 24-hour systolic, nocturnal systolic, and diastolic BP. Higher nicotine dependency scores of FNDT were found to be related to abnormal BP dipping, largely due to higher rates of non-dipper and reverse dipper patterns. Patients with a history of smoking were more likely to have diabetes mellitus, dyslipidemia, and metabolic syndrome.

## Data Availability

All data documents are filed and can be sent upon request. References are given in the manuscript.

## Acknowledgment

We would like to express our sincere gratitude to other medical professionals at our hospital, especially nurse Ayşe Bilgin, for their valuable assistance during the study procedure.

## References

1. Guideline for the pharmacological treatment of hypertension in adults. WHO Guidelines Approved by the Guidelines Review Committee. Geneva2021.

2. Unger T, Borghi C, Charchar F, Khan NA, Poulter NR, Prabhakaran D, et al. 2020 International Society of Hypertension Global Hypertension Practice Guidelines. Hypertension. 2020;75(6):1334-57.

3. Whelton PK, Carey RM, Aronow WS, Casey DE, Jr., Collins KJ, Dennison Himmelfarb C, et al. 2017 ACC/AHA/AAPA/ABC/ACPM/AGS/APhA/ASH/ASPC/NMA/PCNA Guideline for the Prevention, Detection, Evaluation, and Management of High Blood Pressure in Adults: A Report of the American College of Cardiology/American Heart Association Task Force on Clinical Practice Guidelines. Hypertension. 2018;71(6):e13-e115.

4. Williams B, Mancia G, Spiering W, Agabiti Rosei E, Azizi M, Burnier M, et al. 2018 ESC/ESH Guidelines for the management of arterial hypertension. Eur Heart J. 2018;39(33):3021-104.

5. Oktamianti P, Kusuma D, Amir V, Tjandrarini DH, Paramita A. District-Level Inequalities in Hypertension among Adults in Indonesia: A Cross-Sectional Analysis by Sex and Age Group. Int J Environ Res Public Health. 2022;19(20).

6. Collaborators GBDRF. Global, regional, and national comparative risk assessment of 84 behavioural, environmental and occupational, and metabolic risks or clusters of risks for 195 countries and territories, 1990-2017: a systematic analysis for the Global Burden of Disease Study 2017. Lancet. 2018;392(10159):1923-94.

7. Sheikh AB, Sobotka PA, Garg I, Dunn JP, Minhas AMK, Shandhi MMH, et al. Blood Pressure Variability in Clinical Practice: Past, Present and the Future. J Am Heart Assoc. 2023;12(9):e029297.

8. Parati G, Stergiou GS, Dolan E, Bilo G. Blood pressure variability: clinical relevance and application. J Clin Hypertens (Greenwich). 2018;20(7):1133–7.

9. Filippone EJ, Foy AJ, Naccarelli GV. Controversies in Hypertension III: Dipping, Nocturnal Hypertension, and the Morning Surge. Am J Med. 2023;136(7):629–37.

10. Kario K. Nocturnal Hypertension: New Technology and Evidence. Hypertension. 2018;71(6):997–1009.

11. Huart J, Persu A, Lengele JP, Krzesinski JM, Jouret F, Stergiou GS. Pathophysiology of the Nondipping Blood Pressure Pattern. Hypertension. 2023;80(4):719–29.

12. Spallone V. Blood Pressure Variability and Autonomic Dysfunction. Curr Diab Rep. 2018;18(12):137.

13. Hermida RC, Ayala DE, Mojon A, Fernandez JR. Blunted sleep-time relative blood pressure decline increases cardiovascular risk independent of blood pressure level--the “normotensive non-dipper” paradox. Chronobiol Int. 2013;30(1-2):87–98.

14. Kario K, Williams B. Nocturnal Hypertension and Heart Failure: Mechanisms, Evidence, and New Treatments. Hypertension. 2021;78(3):564–77.

15. Gavriilaki M, Anyfanti P, Mastrogiannis K, Gavriilaki E, Lazaridis A, Kimiskidis V, et al. Association between ambulatory blood pressure monitoring patterns with cognitive function and risk of dementia: a systematic review and meta-analysis. Aging Clin Exp Res. 2023;35(4):745–61.

16. Gur M, Elbasan Z, Yildiray Sahin D, Yildiz Koyunsever N, Seker T, Ozaltun B, et al. DNA damage and oxidative status in newly diagnosed, untreated, dipper and non-dipper hypertensive patients. Hypertens Res. 2013;36(2):166–71.

17. Hazukova R, Rezacova M, Pleskot M, Zadak Z, Cermakova E, Taborsky M. DNA damage and arterial hypertension. A systematic review and meta-analysis. Biomed Pap Med Fac Univ Palacky Olomouc Czech Repub. 2024;168(1):15–24.

18. Murray RL, O’Dowd E. Smoking cessation and lung cancer: never too late to quit. Lancet Public Health. 2023;8(9):e664–e5.

19. Cai JA, Zhang YZ, Yu ED, Ding WQ, Li ZS, Zhong L, et al. Association of cigarette smoking with risk of colorectal cancer subtypes classified by gut microbiota. Tob Induc Dis. 2023;21:99.

20. Christenson SA, Smith BM, Bafadhel M, Putcha N. Chronic obstructive pulmonary disease. Lancet. 2022;399(10342):2227–42.

21. Knuuti J, Wijns W, Saraste A, Capodanno D, Barbato E, Funck-Brentano C, et al. 2019 ESC Guidelines for the diagnosis and management of chronic coronary syndromes. Eur Heart J. 2020;41(3):407-77.

22. Fowler CD, Turner JR, Imad Damaj M. Molecular Mechanisms Associated with Nicotine Pharmacology and Dependence. Handb Exp Pharmacol. 2020;258:373–93.

23. Collaborators GBDRF. Global burden of 87 risk factors in 204 countries and territories, 1990-2019: a systematic analysis for the Global Burden of Disease Study 2019. Lancet. 2020;396(10258):1223-49.

24. WHO Global Report on Trends in Prevalence of Tobacco Use 2000-2025. 4th ed. Geneva: World Health Organization; 2021.

25. Brotman DJ, Davidson MB, Boumitri M, Vidt DG. Impaired diurnal blood pressure variation and all-cause mortality. Am J Hypertens. 2008;21(1):92–7.

26. Cai A, Zhong Q, Liu C, Zhou D, Li X, Zhang Y, et al. Associations of systolic and diastolic blood pressure night-to-day ratios with atherosclerotic cardiovascular diseases. Hypertens Res. 2016;39(12):874–8.

27. Borrelli S, Garofalo C, Gabbai FB, Chiodini P, Signoriello S, Paoletti E, et al. Dipping Status, Ambulatory Blood Pressure Control, Cardiovascular Disease, and Kidney Disease Progression: A Multicenter Cohort Study of CKD. Am J Kidney Dis. 2023;81(1):15–24 e1.

28. Gunebakmaz O, Kaya MG, Duran M, Akpek M, Elcik D, Eryol NK. Red blood cell distribution width in ‘non-dippers’ versus ‘dippers’. Cardiology. 2012;123(3):154–9.

29. Palatini P, Reboldi G, Saladini F, Angeli F, Mos L, Rattazzi M, et al. Dipping pattern and short-term blood pressure variability are stronger predictors of cardiovascular events than average 24-h blood pressure in young hypertensive subjects. Eur J Prev Cardiol. 2022;29(10):1377–86.

30. Ozcan F, Turak O, Durak A, Isleyen A, Ucar F, Ginis Z, et al. Red cell distribution width and inflammation in patients with non-dipper hypertension. Blood Press. 2013;22(2):80-5.

31. Sunbul M, Gerin F, Durmus E, Kivrak T, Sari I, Tigen K, et al. Neutrophil to lymphocyte and platelet to lymphocyte ratio in patients with dipper versus non-dipper hypertension. Clin Exp Hypertens. 2014;36(4):217–21.

32. Demir M. The relationship between neutrophil lymphocyte ratio and non-dipper hypertension. Clin Exp Hypertens. 2013;35(8):570–3.

33. de la Sierra A, Redon J, Banegas JR, Segura J, Parati G, Gorostidi M, et al. Prevalence and factors associated with circadian blood pressure patterns in hypertensive patients. Hypertension. 2009;53(3):466–72.

34. Kaya MG, Yarlioglues M, Gunebakmaz O, Gunturk E, Inanc T, Dogan A, et al. Platelet activation and inflammatory response in patients with non-dipper hypertension. Atherosclerosis. 2010;209(1):278–82.

35. Chotruangnapa C, Tansakun T, Roubsanthisuk W. Clinical risk factors and predictive score for the non-dipper profile in hypertensive patients: a case-control study. Clin Hypertens. 2021;27(1):22.

36. Boggia J, Li Y, Thijs L, Hansen TW, Kikuya M, Bjorklund-Bodegard K, et al. Prognostic accuracy of day versus night ambulatory blood pressure: a cohort study. Lancet. 2007;370(9594):1219-29.

37. Charles L, Triscott J, Dobbs B. Secondary Hypertension: Discovering the Underlying Cause. Am Fam Physician. 2017;96(7):453–61.

38. Forshaw PE, Correia ATL, Roden LC, Lambert EV, Rae DE. Sleep characteristics associated with nocturnal blood pressure nondipping in healthy individuals: a systematic review. Blood Press Monit. 2022;27(6):357-70.

39. Hegde S, Ahmed I, Aeddula NR. Secondary Hypertension. StatPearls. Treasure Island (FL)2025.

40. Yang WY, Thijs L, Zhang ZY, Asayama K, Boggia J, Hansen TW, et al. Evidence-based proposal for the number of ambulatory readings required for assessing blood pressure level in research settings: an analysis of the IDACO database. Blood Press. 2018;27(6):341-50.

41. Fagerstrom KO. Measuring degree of physical dependence to tobacco smoking with reference to individualization of treatment. Addict Behav. 1978;3(3-4):235–41.

42. Heatherton TF, Kozlowski LT, Frecker RC, Fagerstrom KO. The Fagerstrom Test for Nicotine Dependence: a revision of the Fagerstrom Tolerance Questionnaire. Br J Addict. 1991;86(9):1119–27.

43. Uysal MA, Kadakal F, Karsidag C, Bayram NG, Uysal O, Yilmaz V. Fagerstrom test for nicotine dependence: reliability in a Turkish sample and factor analysis. Tuberk Toraks. 2004;52(2):115–21.

44. Gilpin EA, Pierce JP, Farkas AJ. Duration of smoking abstinence and success in quitting. J Natl Cancer Inst. 1997;89(8):572–6.

45. Kim SH, Shin C, Kim S, Kim JS, Lim SY, Seo HS, et al. Prevalence of Isolated Nocturnal Hypertension and Development of Arterial Stiffness, Left Ventricular Hypertrophy, and Silent Cerebrovascular Lesions: The KoGES (Korean Genome and Epidemiology Study). J Am Heart Assoc. 2022;11(19):e025641.

46. Bo Y, Kwok KO, Chung VC, Yu CP, Tsoi KK, Wong SY, et al. Short-term reproducibility of ambulatory blood pressure measurements: a systematic review and meta-analysis of 35 observational studies. J Hypertens. 2020;38(11):2095–109.

47. Huh Y, Min Lee C, Cho HJ. Comparison of nicotine dependence between single and multiple tobacco product users among South Korean adults. Tob Induc Dis. 2022;20:22.

48. Verplaetse TL, Weinberger AH, Oberleitner LM, Smith KM, Pittman BP, Shi JM, et al. Effect of doxazosin on stress reactivity and the ability to resist smoking. J Psychopharmacol. 2017;31(7):830–40.

49. Yasuda G, Hasegawa K, Kuji T, Ogawa N, Shimura G, Umemura S, et al. Effects of doxazosin on ambulatory blood pressure and sympathetic nervous activity in hypertensive Type 2 diabetic patients with overt nephropathy. Diabet Med. 2005;22(10):1394–400.

50. Duan L, Li S, Wang L, Jing Y, Li G, Sun Y, et al. Melatonin Plays a Critical Protective Role in Nicotine-Related Abdominal Aortic Aneurysm. Front Physiol. 2020;11:866.

51. Ogedegbe G, Spruill TM, Sarpong DF, Agyemang C, Chaplin W, Pastva A, et al. Correlates of isolated nocturnal hypertension and target organ damage in a population-based cohort of African Americans: the Jackson Heart Study. Am J Hypertens. 2013;26(8):1011–6.

52. Nolde JM, Kiuchi MG, Lugo-Gavidia LM, Ho JK, Chan J, Matthews VB, et al. Nocturnal hypertension: a common phenotype in a tertiary clinical setting associated with increased arterial stiffness and central blood pressure. J Hypertens. 2021;39(2):250–8.

53. Fan HQ, Li Y, Thijs L, Hansen TW, Boggia J, Kikuya M, et al. Prognostic value of isolated nocturnal hypertension on ambulatory measurement in 8711 individuals from 10 populations. J Hypertens. 2010;28(10):2036–45.

54. Fujiwara T, Hoshide S, Kanegae H, Kario K. Cardiovascular Event Risks Associated With Masked Nocturnal Hypertension Defined by Home Blood Pressure Monitoring in the J-HOP Nocturnal Blood Pressure Study. Hypertension. 2020;76(1):259–66.

55. Hermida RC, Crespo JJ, Dominguez-Sardina M, Otero A, Moya A, Rios MT, et al. Bedtime hypertension treatment improves cardiovascular risk reduction: the Hygia Chronotherapy Trial. Eur Heart J. 2020;41(48):4565–76.

56. Hermida RC, Ayala DE, Fernandez JR, Calvo C. Chronotherapy improves blood pressure control and reverts the nondipper pattern in patients with resistant hypertension. Hypertension. 2008;51(1):69–76.

57. Hermida RC, Fernandez JR, Mojon A. Current evidence on the circadian-time-dependent effects of hypertension medications and their combinations in relation to findings of MAPEC and Hygia Chronotherapy Trial. Chronobiol Int. 2020;37(5):751–8.

58. Mackenzie IS, Rogers A, Poulter NR, Williams B, Brown MJ, Webb DJ, et al. Cardiovascular outcomes in adults with hypertension with evening versus morning dosing of usual antihypertensives in the UK (TIME study): a prospective, randomised, open-label, blinded-endpoint clinical trial. Lancet. 2022;400(10361):1417–25.

59. Kjeldsen SE, Egan BM, Narkiewicz K, Kreutz R, Burnier M, Oparil S, et al. TIME to face the reality about evening dosing of antihypertensive drugs in hypertension. Blood Press. 2023;32(1):1-3.

60. Poulter NR, Savopoulos C, Anjum A, Apostolopoulou M, Chapman N, Cross M, et al. Randomized Crossover Trial of the Impact of Morning or Evening Dosing of Antihypertensive Agents on 24-Hour Ambulatory Blood Pressure. Hypertension. 2018;72(4):870–3.

61. Goldman L, Cecil RL, Cooney KA, Elsevier C. Goldman-Cecil medicine. Twenty-seventh edition. ed. Philadelphia, PA: Elsevier; 2024.

62. Maddatu J, Anderson-Baucum E, Evans-Molina C. Smoking and the risk of type 2 diabetes. Transl Res. 2017;184:101–7.

63. Kar D, Gillies C, Zaccardi F, Webb D, Seidu S, Tesfaye S, et al. Relationship of cardiometabolic parameters in non-smokers, current smokers, and quitters in diabetes: a systematic review and meta-analysis. Cardiovasc Diabetol. 2016;15(1):158.

64. Jeong W. Association between dual smoking and dyslipidemia in South Korean adults. PLoS One. 2022;17(7):e0270577.

65. Sun K, Liu J, Ning G. Active smoking and risk of metabolic syndrome: a meta-analysis of prospective studies. PLoS One. 2012;7(10):e47791.

